# Comparison of the prevalence of all diagnosed diseases among Estonian Biobank participants against the general population

**DOI:** 10.64898/2026.02.05.26345634

**Authors:** Maarja Pajusalu, Marek Oja, Kerli Mooses, Silver Heinsar, Oliver Aasmets, Triin Laisk, Priit Palta, Elin Org, Reedik Mägi, Urmo Võsa, Krista Fischer, Estonian Biobank Research Team, Taavi Tillmann, Sven Laur, Sulev Reisberg, Jaak Vilo, Raivo Kolde

## Abstract

Characterizing study sample representativeness is critical for the validity of biobank-derived findings, yet selection biases are rarely quantified across the full clinical spectrum. Here we systematically evaluate the Estonian Biobank (EstBB) - comprising ∼20% of the adult population - by comparing two recruitment waves against a 30% national reference dataset (Est-Health-30). Analyzing prevalence ratios (PR) across 1,028 ICD-10 categories, we reveal a bifurcated landscape of representativeness. While EstBB achieves population parity (PR ∼ 1.0) for widespread chronic conditions like Type 2 diabetes, 47% of diagnostic categories exhibit significant deviations (>1.3-fold). We identify a distinct “managed symptomatic” phenotype: a systematic enrichment of outpatient diagnoses - such as melanocytic nevi (PR=2.07, 95% CI 1.93…2.21) and major depression (PR=1.53, 1.4…1.66) - coupled with a depletion of high-mortality conditions like lung cancer (PR=0.69, 0.64…0.75) and vascular dementia (PR=0.45, 0.38…0.54). These biases evolved across recruitment phases, with the later EstBB2 cohort exhibiting a healthier, prevention-oriented profile. To support research integrity, we provide an interactive open-access dashboard for phenotype refinement. Accounting for such selection-driven “clinical visibility” is essential to avoid collider bias in risk prediction and causal inference models.

## Background

The scientific utility of population-based biobanks is fundamentally linked to their representativeness - the degree to which a cohort reflects the sociodemographic and health characteristics of its target population. While evidence from the UK Biobank suggests that even non-representative cohorts can yield generalizable findings regarding exposure-disease associations, characterizing deviations in disease prevalence remains essential for identifying potential biases. Understanding these differences provides the necessary context for interpreting longitudinal data and evaluating the generalizability of findings derived from the cohort.

Against this background of cohort characterization, the Estonian Biobank (EstBB) provides a unique platform for examining population-wide health trends. Established in 2000, the EstBB has grown to include 212,000 participants as of 2025, representing approximately 20% of Estonia’s adult population. The EstBB dataset provides comprehensive longitudinal integration by linking data from the Estonian Health Insurance Fund (EHIF) and the National Health Information System (NHIS) with centralized medical records dating back to 2004 [1]. Reflecting the value of this integration, the data has been utilized in over 300 research projects and has contributed to more than 800 publications [1]. EstBB’s phenotyping depth has been augmented by specialized initiatives, including personalized medicine pilots for breast cancer and myocardial infarction [2], the Mental Health Online Survey (MHoS) for depression and anxiety [3], and pharmacogenomics research leveraging electronic health records [4]. While these contributions underscore EstBB’s impact on global genomics and personalized medicine [5], the representativeness of its prevalence estimates remains uncharacterized.

Participation in biobanks is influenced by recruitment strategies, socio-demographic factors, and health status, all of which introduce selection bias and alter population representativeness [6], [7]. While public campaigns often induce a “healthy volunteer” effect, hospital-based recruitment leads to disease enrichment [8]. Furthermore, both approaches may be subject to survival bias depending on the temporal window between recruitment and observation. In the case of EstBB, marketing tactics - such as media campaigns targeting younger “digital natives” and pharmacy-based recruitment - have been shown to skew cohort composition [1].

The participant profiles across various biobanks frequently reveal a “healthy-conscious” bias, characterized by overrepresentation of women [9], [10], [11], [12], higher educational attainment [13], [11], and greater trust in institutions [13]. Previous research on the Estonian Biobank (EstBB) aligns with these observations, demonstrating a significant overrepresentation of female participants (66%) [1], [12], [14], [15], a “healthy volunteer bias” characterized by a higher prevalence of university-educated individuals in recent recruitment waves [1], and high institutional trust evidenced by the widespread voluntary participation and adoption of the national digital ID system for secure consent [1].

These health-related biases are non-uniform: while some cohorts report higher baseline prevalence of non-fatal conditions like peptic ulcers, osteoarthritis, or asthma [11], [8], they simultaneously exhibit lower cancer incidence [9] and mortality rates [9], [16] compared to the general population. For instance, the FinnGen study reported significantly higher prevalence for asthma and type 2 diabetes than the population-based FinRegistry, illustrating that even “disease-enriched” cohorts are not strictly representative [8].

While lack of representativeness is not always problematic for some research questions (e.g. quantifying to what extent smoking elevates mortality [16]), it may hamper or invalidate other research questions (e.g. what is the prevalence of a health behaviour, a disease [17], or when creating or recalibrating diagnostic or prognostic clinical risk prediction models). As personalized medicine transitions from discovery to implementation, understanding the exact nature of these biases is paramount. To date, research has largely focused on fatal outcomes or limited sets of conditions. We sought to systematically compare the prevalence of a wide range of diseases against a national reference to facilitate more valid data interpretations.

This study describes and quantifies the differences in the prevalence of diagnoses between the whole Estonian Biobank and the general population. Moreover, the prevalence of diagnoses in two EstBB recruitment phases with different recruitment strategies is compared. We further examined sex-specific disparities by identifying diagnoses that were significantly over- or underrepresented in EstBB, indicating pronounced differences in representation. As a novel approach, an interactive dashboard was developed to provide a detailed overview of all analysed diagnoses and datasets, combining summary tables with interactive visualisations for exploration at multiple granularities.

## Methods

### Data

Four datasets transformed into the Observational Medical Outcomes Partnership Common Data Model (OMOP CDM) were included into the analysis: the EstBB, a representative 30% random sample of the general Estonian population (Est-Health-30), and two subsets of EstBB corresponding to its first and second recruitment phases, labelled as EstBB1 and EstBB2. The first recruitment phase (2004–2010, few more until 2017) enrolled 52,289 participants through a nationwide network of general practitioners, and dedicated recruitment centers, pharmacies and the defence forces. The second recruitment phase between 2018 and 2019, added to the cohort 159,840 participants using large-scale media campaigns and simplified procedures, including online consent forms and self-assessment of some health behaviours and blood sample collection at local healthcare providers or pharmacies (where BMI and blood pressure was usually not collected). The recruitment details are described by Milani et al [1]. The EST-Health-30 dataset provides harmonized OMOP-format health information for 495,000 Estonian residents (population N=1,369,995), healthcare provision claims, electronic health records, lab measurements, prescriptions, cancer, and mortality registries from 2012 to 2024 for a 30% random sample, with updates scheduled through 2026. This sampling fraction was chosen to provide a stable general-population benchmark for stratified analyses. The inclusion criteria for the current analysis were: having a birth year less than or equal to 2005 (age ≥ 18 in 2023); being alive on 1 January 2012; and having at least one prescription or claim between 2012 and 2023. For comparability of case ascertainment across data sources, the analytic cohorts in both Est-Health-30 and EstBB were restricted to individuals with observable healthcare contact during the study period (≥1 reimbursed prescription or insurance claim between 2012 and 2023). We included data from 1 January 2012 to 31 December 2023. Within the Est-Health-30 cohort, this inclusion criterion yields a total of 406,101 individuals and within EstBB 209,985 individuals (EstBB1 50,517, and EstBB2 159,303, missing few with no date of consent). Given that the observation period began in 2012, these youngest participants were a minimum of 10 years old, thereby establishing the 10–19 as the youngest age group included in the study. This work was approved by the Estonian Bioethics and Human Research Council (1.1-12/653, 1.1-12/1039) and the Ethics Committee of University of Tartu (401/T-34).

### Definition of Medical Diagnosis

To compare the prevalence of diagnosis and minimize reporting bias, we extracted the International Classification of Diseases version 10 (ICD-10) diagnosis codes from healthcare provision claims across primary, secondary and tertiary care, as well as from prescriptions. These data sources cover approximately 95% of Estonian population [18]. The raw data were identically collected and transferred to the common data model (OMOP CDM) for both the EstBB and Est-Health-30 datasets [19], [20]. We ran the analysis for ICD-10 category-level diagnosis codes (e.g. F20, Schizophrenia). We excluded the V01–Y98 chapter, which addresses external causes of morbidity as there was an observed data reporting flaw in our workflow for the EstBB cohort between 2012 and 2018 which led to a systematic underestimation of these diagnoses, thus skewing prevalence results before year 2019.

### Disease Prevalence

To provide a comprehensive overview of disease prevalence across the four datasets, we analyzed ICD-10 data, stratified by sex and 10-year age groups. The analysis focused on identifying differences in disease prevalence between datasets. We calculated prevalence (Prev_disease+age+gender+year_) separately for each disease (d), in age group (a: 10–19,, 70–79), and gender (g: M and F) at a given calendar year within the observation period (y 2012-2023). Year of birth was used to determine age. For each demographic group, we calculated prevalence by dividing the number of patients with the diagnosis by the total number of individuals in that group during the year of observation. Gender-age-year groups with fewer than six cases were excluded to ensure statistical reliability and maintain k=5 anonymity [21]. This left us a total of 167,279 prevalence values across 1573 diagnostic groups for Est-Health-30 and 136,497 prevalence values across 1539 diagnostic groups for EstBB.

### Comparison of Disease Prevalence

Prevalence ratios (PR) were calculated as: Prev_dagy_data2/Prev_dagy_data1. To meet the assumptions of the following meta-analysis model, prevalence ratios were log_2_-transformed. This normalization ensures that the distribution of ratios is symmetric around zero, facilitating the aggregation of data across the study period. For interpretability, results were back-transformed to fold differences and reported with 95% confidence intervals (CI) and p-values.

To aggregate estimates across the 12-year study period, we performed a meta-analysis on the log_2_-transformed PRs using the R *meta* package. Yearly estimates for each diagnosis, age, and gender group (PR_dag_) were pooled using a random-effects model with a Paule-Mandel estimator; a fixed-effect model was employed as a fallback if the random-effects model failed to converge [22]. These initial estimates were further aggregated to produce age-combined (PR_dg_) and comprehensive diagnosis-specific (PR_d_) summaries. This left us with 1028 PR_d_ values between EstBB and Est-Health-30. All summary log_2_ estimates and 95% CIs were back-transformed to fold-changes for final interpretation.

### Filtering and Querying Results

To focus the analysis results on meaningful and interpretable differences, we applied filtering criteria to the diagnosis-level prevalence ratios - fold difference in prevalence between datasets above 1.3. A fold difference >1.3 from the perspective of EstBB indicates overrepresentation in the Biobank compared to Est-Health-30, while a fold difference <1/1.3 indicates underrepresentation. We used the years of life lost (YLL) and years lived with disability (YLD) measures to investigate over- and underrepresentation by burden. These data were obtained from the Global Health Estimates 2021, using the Eastern Europe reference table for the year 2021 [23]. These metrics quantify the regional population-level health loss per condition, enabling us to assess if selection biases align with the most burdensome diseases in the target population.

To explore patterns and trends, we developed an interactive dashboard using R and the Shiny package [24], enabling dynamic exploration of diagnosis prevalence available across demographic groups and time (http://omop-apps.cloud.ut.ee/ShinyApps/eh30-estbb-comparison/) [25].

## Results

We quantified diagnostic prevalence differences between the Estonian Biobank (EstBB), its recruitment phases, and the Est-Health-30 population sample (Table 1). Of the 1,535 identified ICD-10 categories, we analyzed 1,028 by calculating prevalence ratios (PR) across demographic strata and aggregating these via meta-analysis. We excluded 507 categories - primarily rare congenital conditions and specific trauma events - due to low event counts. Of the analyzed set, 53% of diagnoses yielded prevalence ratios within the 0.77 ≤ PR ≤ 1.3 range. This subset included widespread chronic conditions such as Type 2 diabetes (PR = 0.99, 95% CI 0.94…1.06) and hypertension (PR = 1.19, 95% CI 1.07…1.33).

**Table 1.**
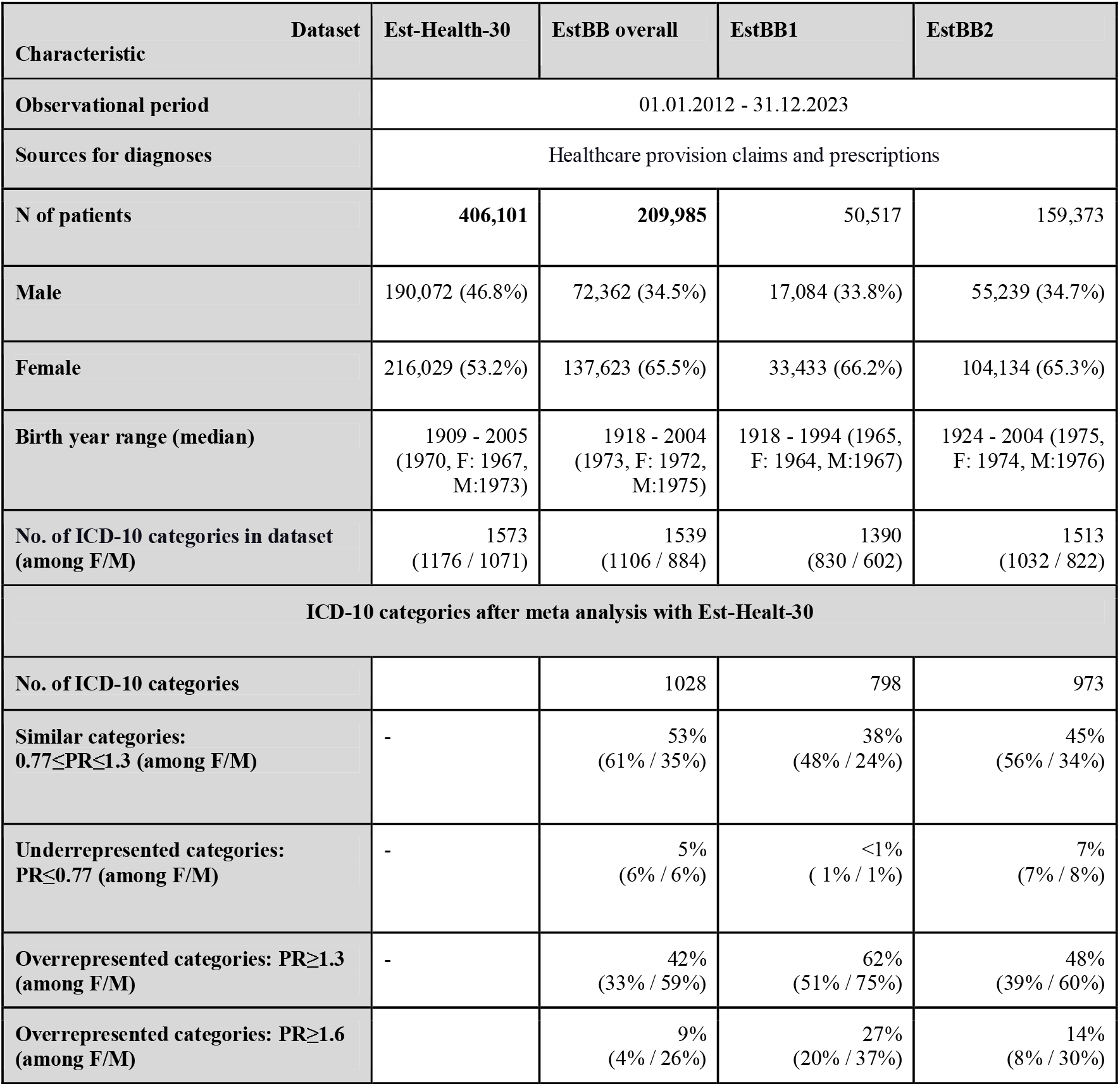
The main characteristics of each dataset.

### Demography of Est-Health-30, EstBB1, EstBB2

To contextualize the observed prevalence differences, we examined the demographic composition of EstBB1, EstBB2, and total EstBB using age-sex distribution, with the Est-Health-30 for reference (Figure 1).

**Figure 1.**
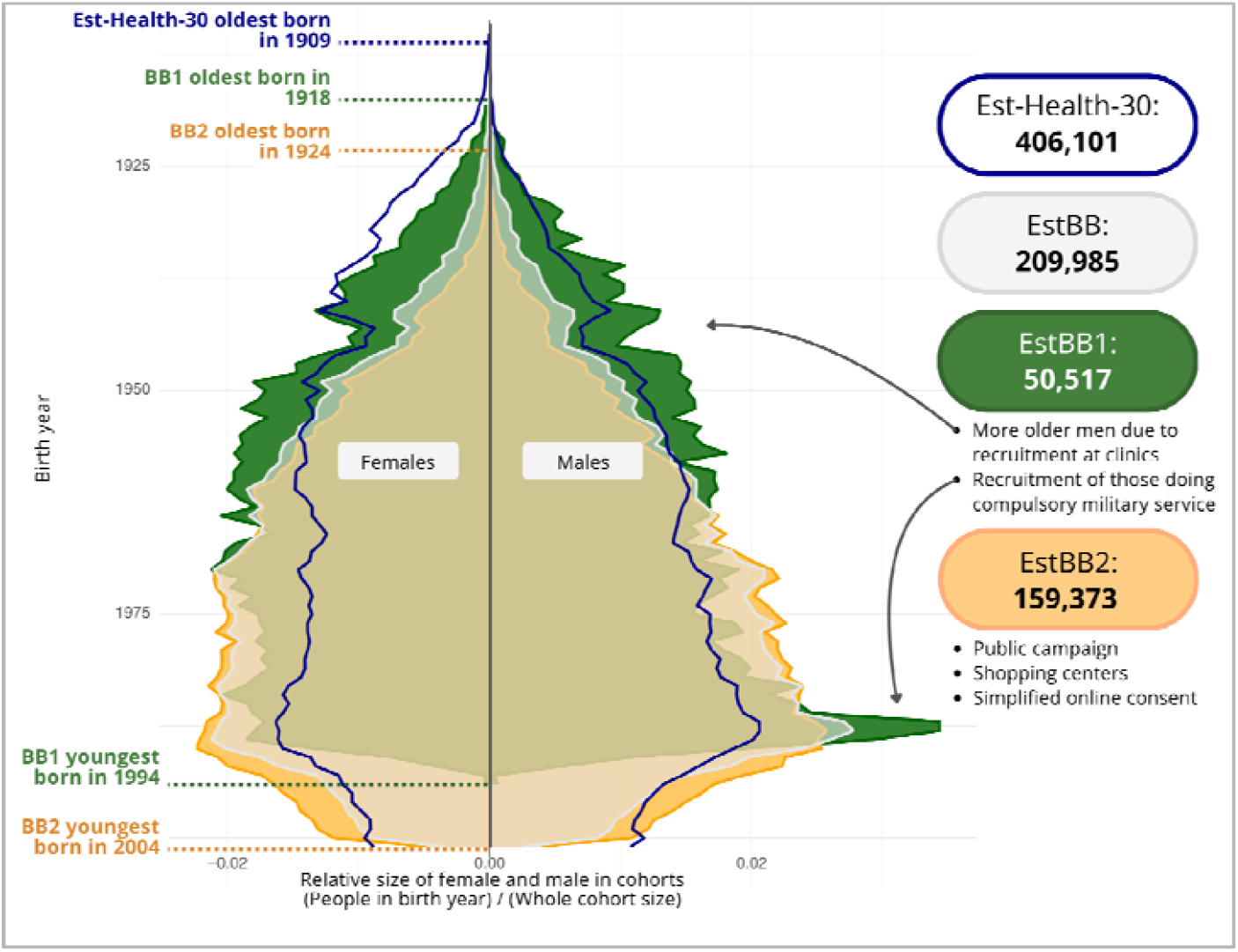
Age and sex distribution of EstBB recruitment waves. Population pyramids for EstBB1 (green), EstBB2 (orange), and the combined EstBB cohort (light gray overlay) compared to the Est-Health-30 (blue line). The x-axis represents the relative proportion of each birth-year group within its respective cohort, stratified by sex (females: left; males: right).

The visualization shows distinct cohort structures shaped by recruitment strategies. In comparison to the Est-Health-30, EstBB1 (main recruitment 2004-2010 and few added until 2017), recruited via general practitioners and medical staff, overrepresents older men while underrepresenting older women. EstBB1 has a relative peak in younger men, particularly those born between 1987-1989 due to recruitment from the military service (around 2005-2006). In contrast, EstBB2 recruited in 2018 and 2019, is skewed toward younger participants, reflecting a healthier and more self-selected population indicating a lower expected disease burden. These demographic imbalances are critical for interpreting prevalence estimates and assessing the representativeness of the biobank data.

Due to demographic constraints, EstBB1 could not be analyzed for childhood diagnoses in the 10-19 age group during 2012-2023. The youngest participants in EstBB1 were born in 1994, making them 18 years old at the start of the observation period. As shown in Figure 1, this cohort is small, further limiting analytical feasibility. From Est-Health-30 we excluded persons born later than 2005 while this was the latest birth year for EstBB.

### Prevalence Differences By Diagnoses, Stratified by Health Burden

Figure 2 illustrates the distribution of prevalence differences for diagnoses between EstBB (2A), EstBB1 (2B), and EstBB2 (2C) in comparison to Est-Health-30 across the observation period. In the combined EstBB cohort, 53% of diagnoses differed by at least 30%, corresponding to PR ≥ 1.3 (overrepresentation) or PR ≤ 0.77 (underrepresentation). Overall, 42% of diagnoses were overrepresented (PR ≥ 1.3) and 5% were underrepresented (PR ≤ 0.77).

**Figure 2.**
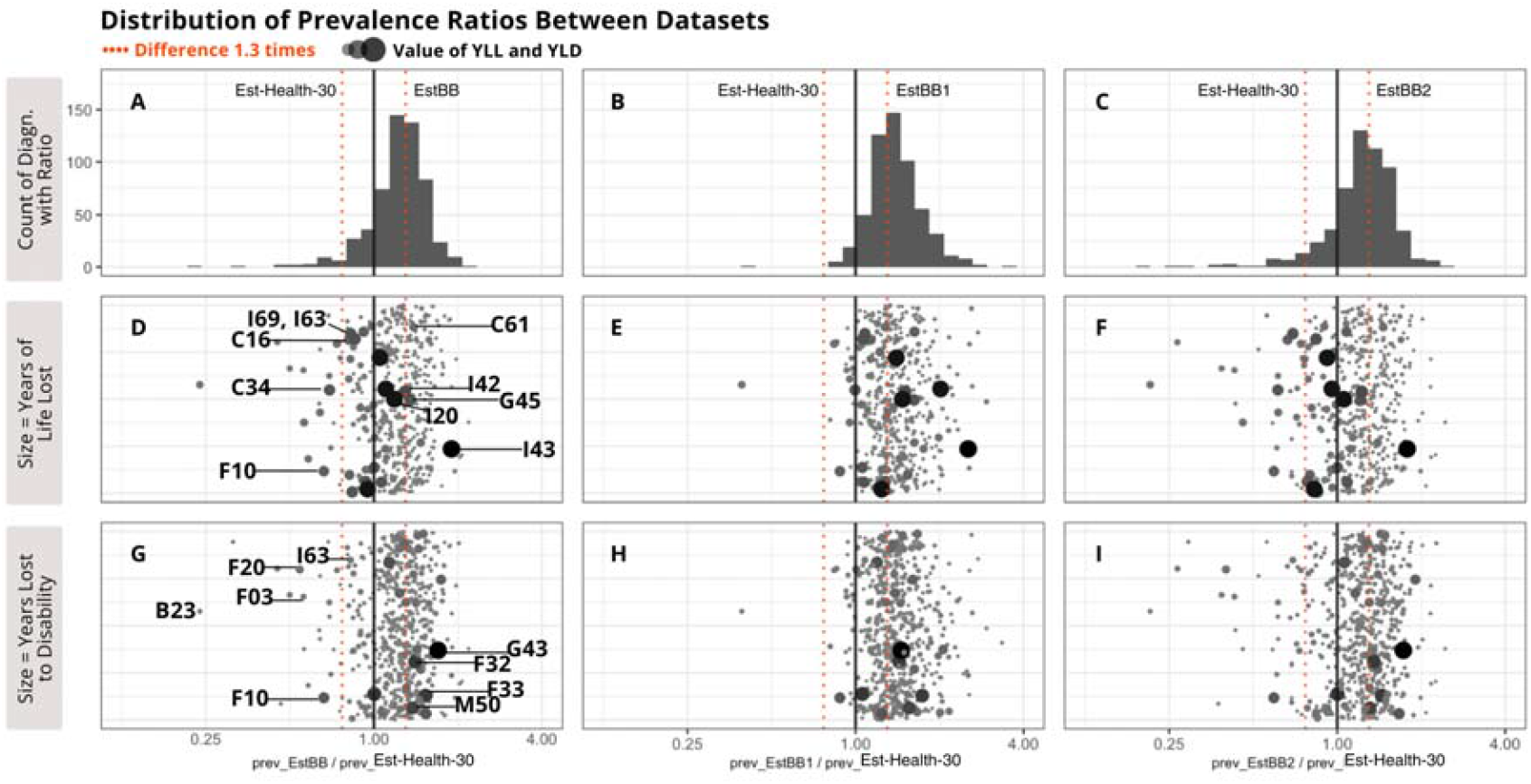
Representation bias in EstBB cohorts by disease burden. Prevalence differences between EstBB, EstBB1, EstBB2, and the general population (Est-Health-30). **Row 1:** Distribution of prevalence differences. **Row 2:** Prevalence differences scaled by Years of Life Lost (YLL). **Row 3:** Prevalence differences scaled by Years Lost to Disability (YLD). Bubble size corresponds to the magnitude of YLL/YLD; the y-axis is jittered for clarity. The red dashed line indicates a 1.3-fold difference threshold.

Differences were more pronounced in the earlier recruitment wave. In EstBB1 (2B), 62% of diagnoses were overrepresented compared with 48% in EstBB2 (2C). Underrepresentation, when applying a fold difference threshold of 1.3, is nearly absent in EstBB1 with only 3 diagnoses related to HIV and immunization compared to 7% in EstBB2 and 5% in combined EstBB.

Utilizing Years of Life Lost (YLL) and Years Lived with Disability (YLD) from the Global Health Estimates 2021 (Eastern Europe reference) [23], we assessed whether selection biases in the Estonian Biobank (EstBB) align with regional population-level health burdens. Our analysis reveals that diagnoses with high YLL are generally underrepresented, whereas those with high YLD are overrepresented (Figure 2D-I).

Across the mortality burden spectrum (Fig. 2D), conditions with high YLL and high lethality—such as lung/stomach cancers (C34, C16) and alcohol-related disorders (F10)—were underrepresented. Conversely, high-YLL conditions associated with chronic clinical management, including angina (I20) and prostate cancer (C61), showed higher prevalence in the EstBB.

In the disability burden analysis (Fig. 2G), we observed a similar trend: underrepresentation was concentrated in conditions involving cognitive or social impairment (e.g., dementias F00-F03), while chronic, non-fatal morbidities like migraine (G43) and major depressive disorder (F32, F33) were overrepresented.

We further evaluated the representativeness of conditions prioritized for international screening and risk-prediction. For instance, prostate cancer (C61) was 1.39-fold overrepresented (PR = 1.39, 95% CI 1.29…1.49), whereas chronic kidney disease (N18) aligned closely with population parity (PR = 1.07, 95% CI 0.94…1.2). Significant depletions were observed for HIV (PR = 0.24, 95% CI 0.18…0.31), opioid-related disorders (PR = 0.19, 95% CI 0.16…0.23), and alcoholic liver disease (PR = 0.56, 95% CI 0.49…0.65), while lung cancer (C34) showed a 1.4-fold underrepresentation (PR = 0.69, 95% CI 0.64…0.75).

### Prevalence Differences By Diagnoses and Sex

Sex-stratified analyses revealed a higher diagnostic burden among males in the EstBB compared to the general population (Figure 3A-C). In the EstBB male cohort, 56% of diagnoses were overrepresented (PR ≥ 1.3), with the highest proportion observed in EstBB1 (71%) followed by EstBB2 (57%).

**Figure 3.**
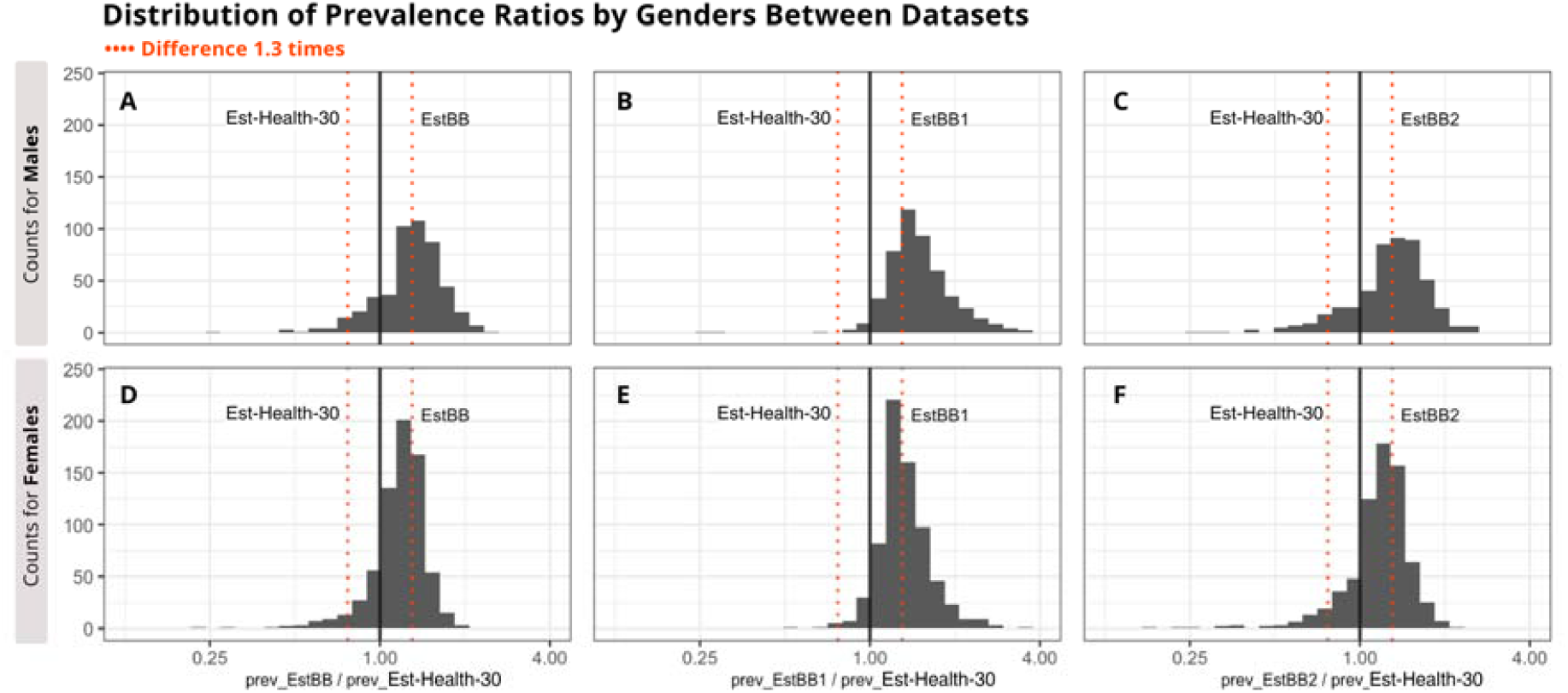
Sex-specific prevalence differences between EstBB cohorts and the general population. Comparison of diagnostic prevalence between EstBB, EstBB1, EstBB2 and the Est-Health-30 reference population, stratified by sex. The red dashed line denotes a 1.3-fold prevalence ratio threshold.

In contrast, female cohorts demonstrated higher representativeness, with prevalence patterns aligning more closely with the general population (Figures 3d-f). The proportion of overrepresented diagnoses (PR ≥ 1.3) was notably lower in females than in males: 30% in the combined EstBB, 47% in EstBB1, and 37% in EstBB2.

Overall, diagnostic overrepresentation was more pervasive and higher in magnitude among males than females across all recruitment waves. While the male sub-cohort exhibited significant deviations from population prevalence across a broad range of diagnoses, the female cohort - characterized by a larger sample size - demonstrated prevalence ratios consistently more similar to the general population.

### Prevalence Ratios by Age and Sex

We computed sex- and age-specific prevalence ratios for all eligible diagnoses. Using ICD-10 chapters F (Mental and behavioural disorders) and I (Diseases of the circulatory system) as examples, Figure 4 illustrates the diagnostic depth of this analysis and the utility of the heatmap tool in identifying age-varying patterns of representation. Other ICD-10 categories can be explored on the interactive dashboard [25].

**Figure 4.**
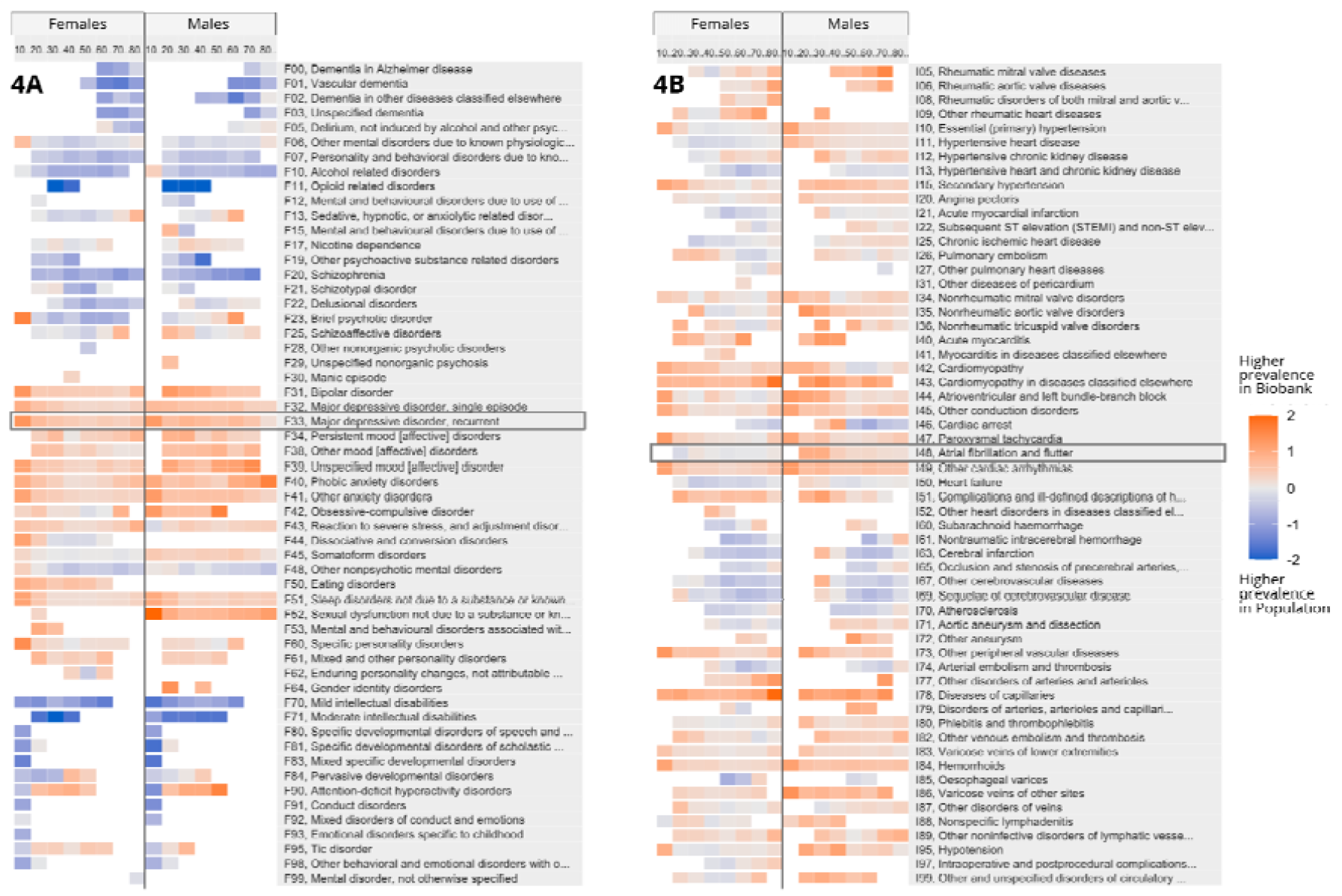
Sex- and age-stratified prevalence ratios for mental and cardiovascular disorders. Comparison of diagnostic prevalence ratios between EstBB and the general population (Est-Health-30) for ICD-10 chapters F (Mental and behavioural disorders) and I (Diseases of the circulatory system). Highlighted points indicate specific diagnoses selected for in-depth analysis.

Within the mental health chapter (Figure 4a), degenerative, dementia-related, and pediatric-onset diagnoses were consistently underrepresented. Conversely, mood and anxiety disorders - specifically depression-related diagnoses - showed overrepresentation. While cardiovascular diseases (Figure 4B) also exhibited widespread deviations from parity (PR ≥ 1.3 or ≤ 0.77), the magnitude of these differences was generally lower.

Comparisons between these chapters highlight that while bias is ubiquitous across both disease classes, the degree of non-representativeness is significantly more pronounced and age-variant in psychiatric disorders than in cardiovascular conditions.

### Prevalence Ratios and Prevalence Values in Detail

While detailed prevalence analyses were performed across all diagnoses [25], we highlight recurrent depressive disorder (F33) and atrial fibrillation and flutter (I48) as representative examples of the interplay between representation bias and absolute prevalence.

Recurrent depressive disorder (F33; Figure 5) was consistently overrepresented across all demographic strata and throughout the observation period. Notably, while the absolute prevalence of mood disorders was higher in females, the representation bias - the magnitude of deviation from parity - was more pronounced among males.

**Figure 5.**
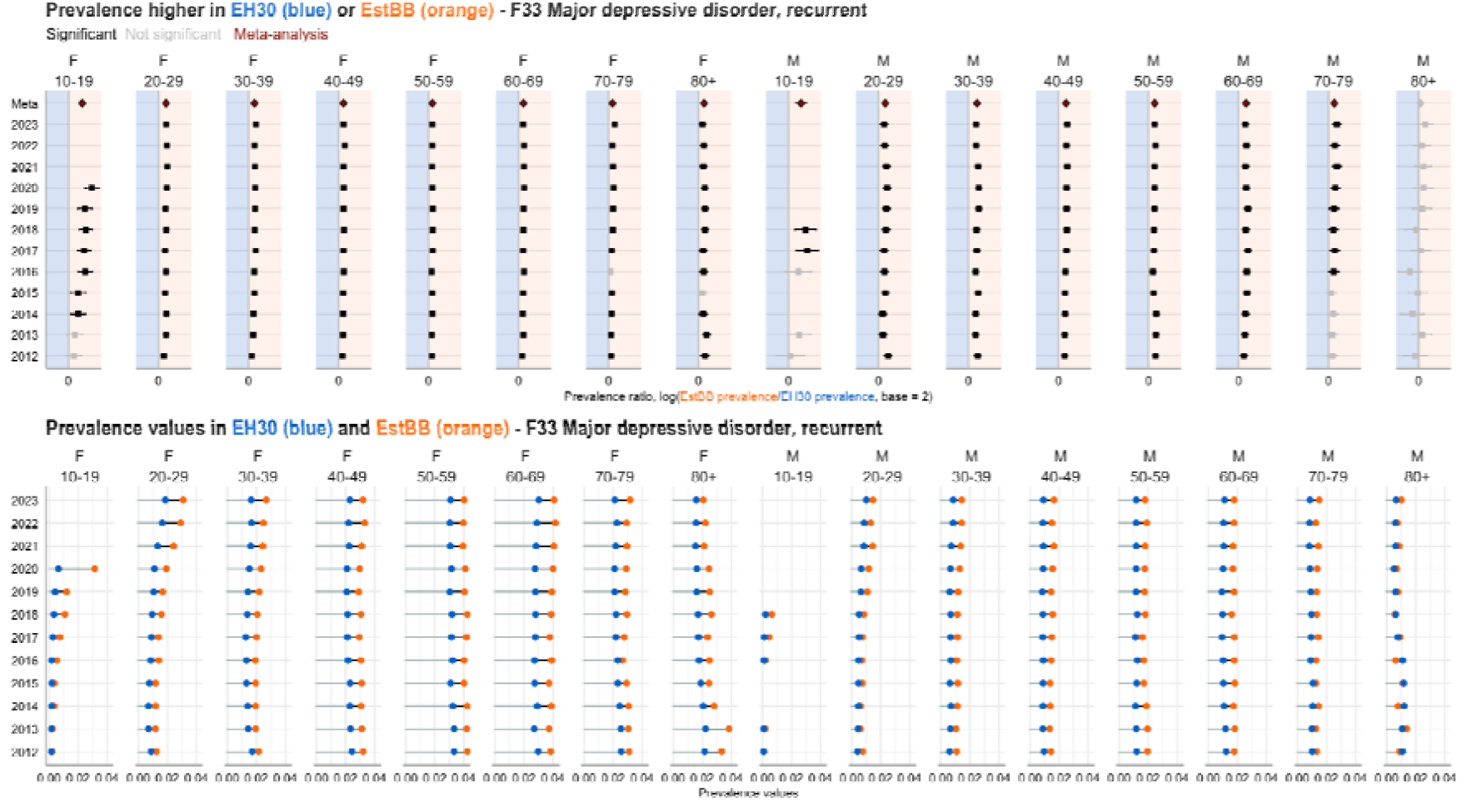
Prevalence ratios with 95% CI and values of recurrent major depressive disorder (F33).

In contrast, the representation of atrial fibrillation and flutter (I48; Figure 6) exhibited a distinct sex-specific pattern. While prevalence in females aligned with population parity, males were consistently overrepresented. This bias was particularly evident in younger males (ages 30–49), who demonstrated disproportionately high PRs despite having low absolute prevalence levels.

**Figure 6.**
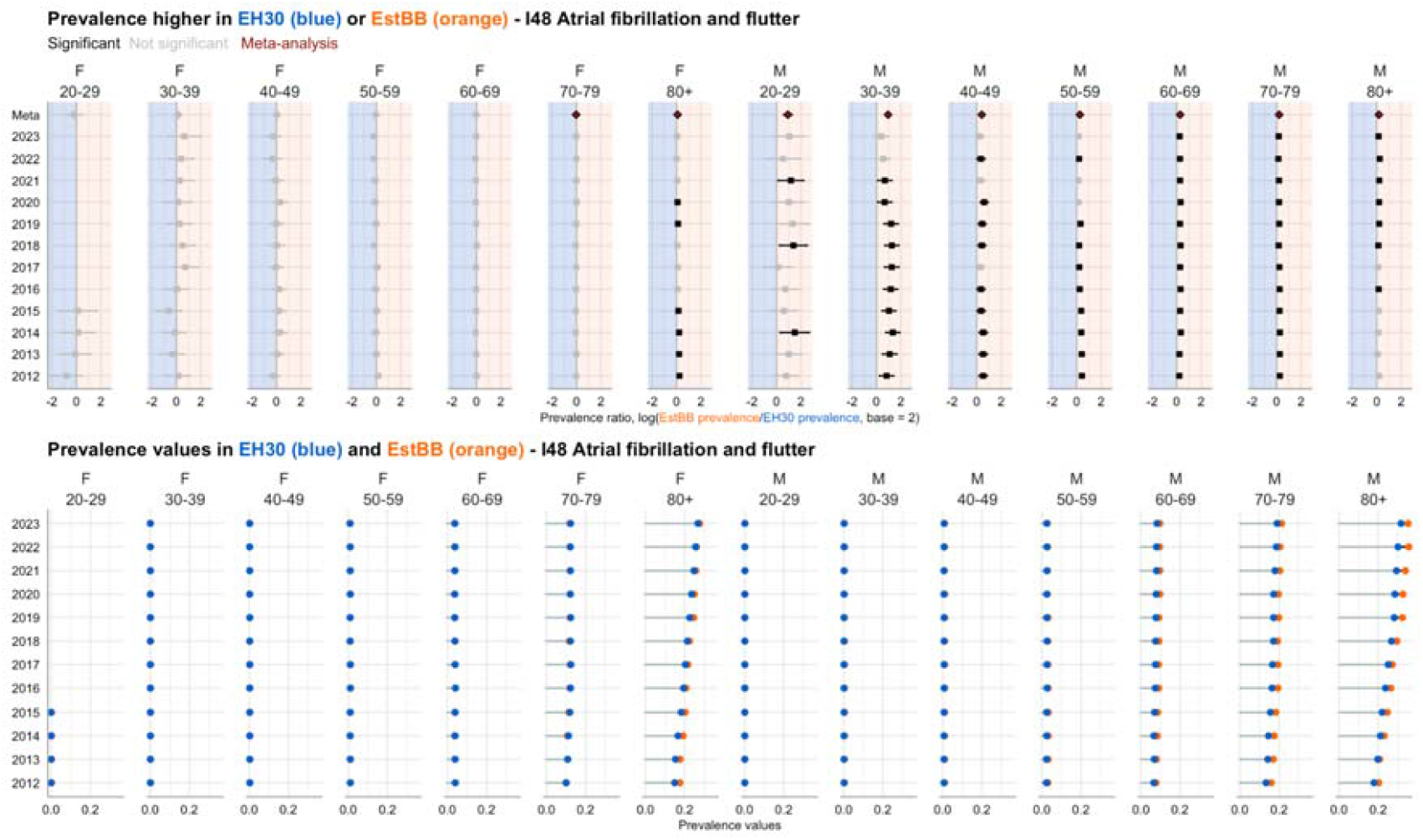
Prevalence ratios and values of atrial fibrillation and flutter (I48).

## Discussion

Selection biases and the “healthy volunteer effect” are well-documented in large-scale biobanks like the UK Biobank, where participants typically exhibit lower rates of smoking, obesity, and severe disease compared to the general public [1], [9], [26]. While the Estonian Biobank (EstBB) is characterized by high nationwide coverage and population capture [1], our analysis of 1,028 diagnostic categories revealed that representativeness remains highly bifurcated. We found that while 53% of diagnoses align with population benchmarks (0.77 ≤ PR ≤ 1.3), the remaining 47% exhibit significant deviations. Specifically, a small number (5%) of high-mortality conditions are underrepresented and a larger proportion (42%) of mostly outpatient and preventive care codes are overrepresented in EstBB. This defines a distinct “managed symptomatic” phenotype: a cohort that is not merely “healthier,” but more proactively engaged with clinical surveillance. Despite these shifts, the EstBB maintains high population parity (PR ∼ 1.0) for high-burden chronic conditions like Type 2 diabetes and hypertension, confirming its validity for epidemiological modeling in these domains.

The observed divergence between mortality (YLL) and disability (YLD) burdens highlight the “managed symptomatic” phenotype. Participants in this niche are stable enough to bypass the attrition bias seen in high-mortality conditions (e.g., lung cancer, C34), yet sufficiently engaged with healthcare to have conditions like hypothyroidism (E03) or angina (I20) documented at earlier stages than the general population. These effects are magnified among males, displaying for example a higher diagnostic in urogenital and neoplastic conditions, suggesting motivation by specific medical concerns. In contrast, the female cohort demonstrates significantly closer alignment with population parity, potentially reflecting the better health engagement of the female population in general.

The two waves of recruitment add a layer of complexity. The earlier EstBB1 cohort (2004–2010) exhibits a higher disease burden and greater socio-economic diversity, including higher rates of smoking and lower educational attainment, while the EstBB2 cohort (2018–2019) is characterized by higher health literacy [1]. Such differences are unavoidable, when recruiting large population based volunteer cohorts, but have to be taken into account for effective use of the cohort. Whether to use a pooled cohort, the two separately or only one, depends very much on the goals of planned study.

The current study adds a layer of detail to account for making such decisions. We examine differences between cohorts and also the general population in terms of diagnosis prevalence very granularly, on a single diagnosis, age group and year level. To enable navigation of researchers to navigate these results, we developed an open-access diagnostic dashboard (http://omop-apps.cloud.ut.ee/ShinyApps/eh30-estbb-comparison), providing age-stratified PR heatmaps and raw data for all 1,028 compared diagnoses. This facilitates phenotype refinement, determination of the study periods, selection of the subcohorts and evaluating the effect of diagnostic changes across years. Also, this information is crucial for interpreting the results for a particular study in terms of potential biases, such as selection, survival and even collider bias [27].

### Strengths and Limitations

Some limitations must be considered. First, the EstBB1 cohort lacks data on participants under age 20 due to its 18-year recruitment minimum (2004–2010), limiting bias inferences for younger populations. Second, the use of three-character ICD-10 categories - while necessary for broad comparison - masks clinical heterogeneity such as disease severity or stage. Third, the exclusion of rare diagnoses (<6 cases) to maintain k-anonymity and statistical stability reduces the power to detect selection bias in low-frequency, high-impact conditions. Finally, despite using administrative data to mitigate reporting bias, unmeasured variations in healthcare-seeking behavior or regional coding practices may confound certain prevalence estimates.

A primary strength of the study is the use of Est-Health-30, a 30% representative sample of the general population, providing a robust comparator. Moreover, methodological consistency was secured by transforming all datasets into the OMOP Common Data Model with the same workflow, ensuring that diagnoses for both cohorts were derived from identical sources and using the same principles. Furthermore, the application of random-effects meta-analysis across 12 years of data effectively accounted for temporal heterogeneity. The granularity of our sex- and age-stratified approach provided sufficient power to identify recruitment-specific disparities (EstBB1 vs. EstBB2). Finally, as a novel approach, we developed the R/Shiny dashboard which offers a transparent, dynamic resource for the research community, facilitating hypothesis generation and data exploration beyond the constraints of a static publication.

## Conclusions

The Estonian Biobank (EstBB) offers a valuable resource for studying genetic underpinnings of a variety of health conditions, supported by detailed phenotyping and longitudinal data. However, as volunteer based biobanks, it does not fully represent the general Estonian population for all health conditions. Although more than half of its diagnoses exhibit similar prevalence values to the general population, its limited representation of some severe, late-stage, and high-mortality diseases - such as advanced cancers and dementia, respiratory failure, - reflects healthy volunteer bias and targeted recruitment decisions. These factors have to be taken into account when considering its representativeness for population-level disease burden estimations. To support research integrity, we provide an interactive open-access dashboard for phenotype refinement.

## Supporting information

Supplementary 1. PR values per diagnosis in EstBB in comparison to Est-Health-30.

## Data availability statement

The data analyzed in this study was obtained from the Estonian Biobank (EstBB). Individual level data from the Estonian Biobank or Est-Health-30 cannot be shared publicly due to legal and ethical restrictions. According to the Human Genes Research Act, which regulates the operations of the Estonian Biobank, the data include potentially identifiable genetic and health information and are therefore subject to restricted access. Access to these data is only possible directly through the Estonian Biobank upon ethics approval from the Estonian Committee on Bioethics and Human Research. Researchers may request access following the procedure described here: https://genomics.ut.ee/en/content/estonian-biobank#dataaccess and requests should be directed to.

The data generated and used as aggregated data for the analysis can be accessed via the open-access diagnostic dashboard developed during this study (http://omop-apps.cloud.ut.ee/ShinyApps/eh30-estbb-comparison) or contacting the authors directly.

## Ethics approval

This work was approved by the Estonian Bioethics and Human Research Council (1.1-12/653, 1.1-12/1039) and the Ethics Committee of University of Tartu (401/T-34).

## Acknowledgments

We acknowledge the Estonian Biobank research team: Andres Metspalu, Lili Milani, Tõnu Esko, Mait Metspalu, giving them credit for data collection, genotyping, QC, and imputation.

## Author contributions

MP: Methodology, Data curation, Investigation, Resources, Visualization, Writing – original draft, Writing – review & editing. MO: Data curation, Methodology, Writing – review. KM: Supervision, Writing – review & editing. SH: Investigation, Writing – review & editing. OA: Writing – review & editing. TL: Writing – review & editing. PP: Writing – review & editing. EO: Writing – review & editing. RM: Conceptualization, Methodology, Writing – review & editing. UV: Conceptualization. KF: Methodology, Writing – review & editing. Estonian Biobank Research Team: Data curation, Investigation, Resources. TT: Conceptualization, Methodology, Writing – review & editing. SL: Writing – review & editing. SR: Writing – review & editing. JV: Funding acquisition, Resources, Writing – review & editing. RK: Conceptualization, Methodology, Funding acquisition, Investigation, Methodology, Resources, Supervision, Writing – review & editing.

