## Supplementary 1. PR values per diagnosis in EstBB in comparison to Est-Health-30. for "Comparison of the prevalence of all diagnosed diseases among Estonian Biobank participants against the general population"

Showing PR values PR<0.77 (underrepresented in EstBB; blue) or PR>1.3 (overrepresented in EstBB; orange) and p-value<0.05.

| **ICD-10** | **Diagnosis category** | **PR (95% CI)** | **P-val** |
| --- | --- | --- | --- |
| A41 | Other sepsis | 0.68 (0.63…0.74) | <0.001 |
| A54 | Gonococcal infection | 1.49 (1.24…1.79) | <0.001 |
| A56 | Other sexually transmitted chlamydial diseases | 1.34 (1.21…1.49) | <0.001 |
| A60 | Anogenital herpesviral [herpes simplex] infections | 1.63 (1.49…1.8) | <0.001 |
| A63 | Other predominantly sexually transmitted diseases, not elsewhere classified | 1.41 (1.26…1.57) | <0.001 |
| A69 | Other spirochetal infections | 1.67 (1.56…1.79) | <0.001 |
| A74 | Other diseases caused by chlamydiae | 1.42 (1.27…1.57) | <0.001 |
| A84 | Tick-borne viral encephalitis | 1.45 (1.08…1.94) | 0.01 |
| B00 | Herpesviral [herpes simplex] infections | 1.66 (1.56…1.76) | <0.001 |
| B07 | Viral warts | 1.67 (1.53…1.82) | <0.001 |
| B20 | Human immunodeficiency virus [HIV] disease resulting in infectious and parasitic diseases | 0.31 (0.19…0.51) | <0.001 |
| B23 | Human immunodeficiency virus [HIV] disease resulting in other conditions | 0.24 (0.18…0.31) | <0.001 |
| B27 | Infectious mononucleosis | 1.42 (1.31…1.55) | <0.001 |
| B33 | Other viral diseases, not elsewhere classified | 1.34 (1.26…1.42) | <0.001 |
| B35 | Dermatophytosis | 1.42 (1.34…1.5) | <0.001 |
| B36 | Other superficial mycoses | 1.45 (1.36…1.55) | <0.001 |
| B37 | Candidiasis | 1.4 (1.31…1.49) | <0.001 |
| B49 | Unspecified mycosis | 1.35 (1…1.82) | 0.05 |
| B50 | Plasmodium falciparum malaria | 1.67 (1.25…2.24) | <0.001 |
| B80 | Enterobiasis | 1.38 (1.21…1.59) | <0.001 |
| C17 | Malignant neoplasm of small intestine | 1.56 (1.1…2.21) | 0.01 |
| C32 | Malignant neoplasm of larynx | 0.62 (0.47…0.82) | <0.001 |
| C34 | Malignant neoplasm of bronchus and lung | 0.69 (0.64…0.75) | <0.001 |
| C43 | Malignant melanoma of skin | 1.57 (1.42…1.75) | <0.001 |
| C44 | Other and unspecified malignant neoplasm of skin | 1.53 (1.45…1.61) | <0.001 |
| C57 | Malignant neoplasm of other and unspecified female genital organs | 1.46 (1…2.12) | 0.05 |
| C61 | Malignant neoplasm of prostate | 1.39 (1.29…1.49) | <0.001 |
| C62 | Malignant neoplasm of testis | 1.6 (1.21…2.12) | 0.001 |
| C65 | Malignant neoplasm of renal pelvis | 1.98 (1.39…2.81) | <0.001 |
| C73 | Malignant neoplasm of thyroid gland | 1.6 (1.45…1.76) | <0.001 |
| C81 | Hodgkin lymphoma | 1.45 (1.32…1.59) | <0.001 |
| C82 | Follicular lymphoma | 1.6 (1.22…2.11) | <0.001 |
| C83 | Non-Hodgkin lymphoma | 1.42 (1.25…1.62) | <0.001 |
| C85 | Other and unspecified non-Hodgkin lymphoma | 1.69 (1.11…2.58) | 0.01 |
| C91 | Lymphoid leukemia | 1.44 (1.29…1.61) | <0.001 |
| C92 | Myeloid leukaemia | 1.47 (1.14…1.9) | 0.003 |
| D03 | Melanoma in situ | 1.69 (1.38…2.08) | <0.001 |
| D04 | Carcinoma in situ of skin | 1.75 (1.62…1.9) | <0.001 |
| D05 | Carcinoma in situ of breast | 1.63 (1.33…2.01) | <0.001 |
| D07 | Carcinoma in situ of other and unspecified sites | 1.43 (1…2.03) | 0.05 |
| D10 | Benign neoplasm of mouth and pharynx | 1.48 (1.33…1.66) | <0.001 |
| D12 | Benign neoplasm of colon, rectum, anus and anal canal | 1.36 (1.27…1.46) | <0.001 |
| D14 | Benign neoplasm of middle ear and respiratory system | 1.3 (1.19…1.41) | <0.001 |
| D16 | Benign neoplasm of bone and articular cartilage | 1.4 (1.28…1.54) | <0.001 |
| D17 | Benign lipomatous neoplasm | 1.33 (1.23…1.44) | <0.001 |
| D18 | Hemangioma and lymphangioma any site | 1.74 (1.6…1.89) | <0.001 |
| D21 | Other benign neoplasms of connective and other soft tissue | 1.37 (1.32…1.44) | <0.001 |
| D22 | Melanocytic nevi | 2.07 (1.93…2.21) | <0.001 |
| D23 | Other benign neoplasms of skin | 1.63 (1.55…1.72) | <0.001 |
| D29 | Benign neoplasm of male genital organs | 2.05 (1.73…2.43) | <0.001 |
| D31 | Benign neoplasm of eye and adnexa | 1.4 (1.25…1.58) | <0.001 |
| D35 | Benign neoplasm of other and unspecified endocrine glands | 1.35 (1.15…1.58) | <0.001 |
| D40 | Neoplasm of uncertain or unknown behaviour of male genital organs | 1.5 (1.18…1.92) | 0.001 |
| D47 | Other neoplasms of uncertain behavior of lymphoid hematopoietic and related tissue | 1.34 (1.23…1.46) | <0.001 |
| D48 | Neoplasm of uncertain behavior of other and unspecified sites | 1.62 (1.5…1.74) | <0.001 |
| D68 | Other coagulation defects | 1.53 (1.42…1.65) | <0.001 |
| D72 | Other disorders of white blood cells | 1.64 (1.46…1.85) | <0.001 |
| D80 | Immunodeficiencies with predominantly antibody defects | 1.42 (1.08…1.87) | 0.01 |
| D86 | Sarcoidosis | 1.5 (1.35…1.67) | <0.001 |
| D89 | Other disorders involving the immune mechanism, not elsewhere classified | 1.39 (1.21…1.59) | <0.001 |
| E01 | Iodine-deficiency-related thyroid disorders and allied conditions | 0.73 (0.57…0.96) | 0.02 |
| E04 | Other nontoxic goiter | 1.31 (1.23…1.41) | <0.001 |
| E05 | Thyrotoxicosis [hyperthyroidism] | 1.34 (1.22…1.48) | <0.001 |
| E06 | Thyroiditis | 1.36 (1.28…1.44) | <0.001 |
| E12 | Malnutrition-related diabetes mellitus | 1.34 (1…1.81) | 0.05 |
| E22 | Hyperfunction of pituitary gland | 1.31 (1.11…1.54) | 0.001 |
| E23 | Hypofunction and other disorders of the pituitary gland | 1.39 (1.15…1.68) | <0.001 |
| E27 | Other disorders of adrenal gland | 1.53 (1.24…1.89) | <0.001 |
| E29 | Testicular dysfunction | 2.03 (1.78…2.32) | <0.001 |
| E61 | Deficiency of other nutrient elements | 1.46 (1.22…1.75) | <0.001 |
| E72 | Other disorders of amino-acid metabolism | 1.47 (1.04…2.08) | 0.03 |
| E73 | Lactose intolerance | 1.54 (1.41…1.68) | <0.001 |
| E74 | Other disorders of carbohydrate metabolism | 1.46 (1.35…1.58) | <0.001 |
| E80 | Disorders of porphyrin and bilirubin metabolism | 1.44 (1.32…1.57) | <0.001 |
| E83 | Disorders of mineral metabolism | 1.37 (1.22…1.54) | <0.001 |
| E86 | Volume depletion | 1.46 (1.03…2.08) | 0.03 |
| E87 | Other disorders of fluid, electrolyte and acid-base balance | 0.76 (0.72…0.79) | <0.001 |
| E89 | Postprocedural endocrine and metabolic complications and disorders, not elsewhere classified | 1.52 (1.4…1.66) | <0.001 |
| F00 | Dementia in Alzheimer disease | 0.69 (0.54…0.88) | 0.003 |
| F01 | Vascular dementia | 0.45 (0.38…0.54) | <0.001 |
| F02 | Dementia in other diseases classified elsewhere | 0.58 (0.47…0.72) | <0.001 |
| F03 | Unspecified dementia | 0.56 (0.47…0.67) | <0.001 |
| F07 | Personality and behavioral disorders due to known physiological condition | 0.69 (0.65…0.72) | <0.001 |
| F10 | Alcohol related disorders | 0.66 (0.59…0.74) | <0.001 |
| F11 | Opioid related disorders | 0.19 (0.16…0.23) | <0.001 |
| F19 | Other psychoactive substance related disorders | 0.58 (0.44…0.77) | <0.001 |
| F20 | Schizophrenia | 0.54 (0.5…0.59) | <0.001 |
| F22 | Delusional disorders | 0.76 (0.66…0.88) | <0.001 |
| F31 | Bipolar disorder | 1.6 (1.43…1.79) | <0.001 |
| F32 | Major depressive disorder, single episode | 1.41 (1.32…1.5) | <0.001 |
| F33 | Major depressive disorder, recurrent | 1.53 (1.41…1.66) | <0.001 |
| F34 | Persistent mood [affective] disorders | 1.42 (1.29…1.57) | <0.001 |
| F38 | Other mood [affective] disorders | 1.38 (1.26…1.51) | <0.001 |
| F39 | Unspecified mood [affective] disorder | 1.5 (1.35…1.66) | <0.001 |
| F40 | Phobic anxiety disorders | 1.5 (1.36…1.66) | <0.001 |
| F41 | Other anxiety disorders | 1.45 (1.32…1.58) | <0.001 |
| F42 | Obsessive-compulsive disorder | 1.44 (1.18…1.74) | <0.001 |
| F43 | Reaction to severe stress, and adjustment disorders | 1.31 (1.23…1.39) | <0.001 |
| F50 | Eating disorders | 1.59 (1.44…1.76) | <0.001 |
| F51 | Sleep disorders not due to a substance or known physiological condition | 1.31 (1.22…1.41) | <0.001 |
| F52 | Sexual dysfunction not due to a substance or known physiological condition | 2.02 (1.56…2.61) | <0.001 |
| F53 | Mental and behavioural disorders associated with the puerperium, not elsewhere classified | 1.67 (1.35…2.06) | <0.001 |
| F61 | Mixed and other personality disorders | 1.36 (1.26…1.47) | <0.001 |
| F64 | Gender identity disorders | 2.13 (1.44…3.15) | <0.001 |
| F70 | Mild intellectual disabilities | 0.46 (0.42…0.52) | <0.001 |
| F71 | Moderate intellectual disabilities | 0.31 (0.28…0.35) | <0.001 |
| F80 | Specific developmental disorders of speech and language | 0.5 (0.37…0.67) | <0.001 |
| F83 | Mixed specific developmental disorders | 0.34 (0.27…0.44) | <0.001 |
| F90 | Attention-deficit hyperactivity disorders | 1.44 (1.06…1.97) | 0.02 |
| F91 | Conduct disorders | 0.48 (0.38…0.61) | <0.001 |
| F93 | Emotional disorders specific to childhood | 0.57 (0.35…0.94) | 0.03 |
| F98 | Other behavioral and emotional disorders with onset usually occurring in childhood and adolescence | 0.71 (0.51…0.99) | 0.04 |
| G25 | Other extrapyramidal and movement disorders | 1.58 (1.44…1.73) | <0.001 |
| G31 | Other degenerative diseases of nervous system, not elsewhere classified | 0.5 (0.4…0.63) | <0.001 |
| G35 | Multiple sclerosis | 1.4 (1.25…1.57) | <0.001 |
| G43 | Migraine | 1.7 (1.56…1.85) | <0.001 |
| G44 | Other headache syndromes | 1.3 (1.23…1.38) | <0.001 |
| G45 | Transient cerebral ischemic attacks and related syndromes | 1.35 (1.29…1.41) | <0.001 |
| G47 | Sleep disorders | 1.3 (1.2…1.4) | <0.001 |
| G50 | Disorders of trigeminal nerve | 1.39 (1.28…1.52) | <0.001 |
| G54 | Nerve root and plexus disorders | 1.46 (1.32…1.62) | <0.001 |
| G55 | Nerve root and plexus compressions in diseases classified elsewhere | 1.56 (1.22…1.99) | <0.001 |
| G56 | Mononeuropathies of upper limb | 1.33 (1.21…1.45) | <0.001 |
| G57 | Mononeuropathies of lower limb | 1.35 (1.26…1.45) | <0.001 |
| G58 | Other mononeuropathies | 1.34 (1.21…1.47) | <0.001 |
| G61 | Inflammatory polyneuropathy | 1.45 (1.01…2.07) | 0.04 |
| G70 | Myasthenia gravis and other neuromuscular junction disorders | 1.59 (1.39…1.81) | <0.001 |
| G72 | Other myopathies | 1.56 (1.33…1.82) | <0.001 |
| G80 | Cerebral palsy | 0.61 (0.56…0.66) | <0.001 |
| G81 | Hemiplegia and hemiparesis | 0.64 (0.57…0.72) | <0.001 |
| H00 | Hordeolum and chalazion | 1.37 (1.29…1.44) | <0.001 |
| H01 | Other inflammation of eyelid | 1.51 (1.44…1.58) | <0.001 |
| H02 | Other disorders of eyelid | 1.38 (1.34…1.42) | <0.001 |
| H04 | Disorders of lacrimal system | 1.41 (1.34…1.49) | <0.001 |
| H10 | Conjunctivitis | 1.34 (1.3…1.37) | <0.001 |
| H11 | Other disorders of conjunctiva | 1.31 (1.24…1.37) | <0.001 |
| H18 | Other disorders of cornea | 1.51 (1.38…1.65) | <0.001 |
| H20 | Iridocyclitis | 1.31 (1.2…1.43) | <0.001 |
| H21 | Other disorders of iris and ciliary body | 1.65 (1.27…2.14) | <0.001 |
| H26 | Other cataract | 1.32 (1.24…1.4) | <0.001 |
| H31 | Other disorders of choroid | 1.63 (1.26…2.1) | <0.001 |
| H33 | Retinal detachments and breaks | 1.45 (1.32…1.6) | <0.001 |
| H43 | Disorders of vitreous body | 1.46 (1.39…1.55) | <0.001 |
| H49 | Paralytic strabismus | 1.42 (1.11…1.82) | 0.005 |
| H50 | Other disorders of binocular movement | 1.34 (1.16…1.56) | <0.001 |
| H52 | Disorders of refraction and accommodation | 1.41 (1.34…1.49) | <0.001 |
| H53 | Visual disturbances | 1.37 (1.29…1.46) | <0.001 |
| H61 | Other disorders of external ear | 1.33 (1.29…1.37) | <0.001 |
| H62 | Disorders of external ear in diseases classified elsewhere | 1.41 (1.3…1.53) | <0.001 |
| H69 | Other and unspecified disorders of Eustachian tube | 1.52 (1.42…1.63) | <0.001 |
| H71 | Cholesteatoma of middle ear | 1.48 (1.07…2.04) | 0.02 |
| H72 | Perforation of tympanic membrane | 1.32 (1.17…1.49) | <0.001 |
| H74 | Other disorders of middle ear mastoid | 1.34 (1.19…1.51) | <0.001 |
| H80 | Otosclerosis | 1.34 (1.21…1.49) | <0.001 |
| H82 | Vertiginous syndromes in diseases classified elsewhere | 1.32 (1.11…1.56) | 0.001 |
| H83 | Other disorders of inner ear | 1.47 (1.19…1.81) | <0.001 |
| H92 | Otalgia and effusion of ear | 1.32 (1.26…1.39) | <0.001 |
| H93 | Other disorders of ear, not elsewhere classified | 1.49 (1.35…1.63) | <0.001 |
| H95 | Postprocedural disorders of ear and mastoid process, not elsewhere classified | 1.58 (1.19…2.11) | 0.002 |
| I05 | Rheumatic mitral valve diseases | 1.42 (1.15…1.74) | <0.001 |
| I06 | Rheumatic aortic valve diseases | 1.42 (1.18…1.73) | <0.001 |
| I08 | Rheumatic disorders of both mitral and aortic valves | 1.32 (1.04…1.69) | 0.02 |
| I09 | Other rheumatic heart diseases | 1.43 (1.13…1.82) | 0.003 |
| I15 | Secondary hypertension | 1.36 (1.23…1.5) | <0.001 |
| I40 | Acute myocarditis | 1.6 (1.33…1.92) | <0.001 |
| I41 | Myocarditis in diseases classified elsewhere | 1.34 (1…1.8) | 0.05 |
| I43 | Cardiomyopathy in diseases classified elsewhere | 1.9 (1.71…2.11) | <0.001 |
| I45 | Other conduction disorders | 1.36 (1.25…1.48) | <0.001 |
| I49 | Other cardiac arrhythmias | 1.51 (1.38…1.65) | <0.001 |
| I51 | Complications and ill-defined descriptions of heart disease | 1.51 (1.36…1.67) | <0.001 |
| I52 | Other heart disorders in diseases classified elsewhere | 1.41 (1.06…1.87) | 0.02 |
| I72 | Other aneurysm | 1.44 (1.22…1.7) | <0.001 |
| I73 | Other peripheral vascular diseases | 1.46 (1.32…1.62) | <0.001 |
| I77 | Other disorders of arteries and arterioles | 1.65 (1.3…2.1) | <0.001 |
| I78 | Diseases of capillaries | 1.95 (1.81…2.1) | <0.001 |
| I84 | Hemorrhoids | 1.42 (1.31…1.54) | <0.001 |
| I86 | Varicose veins of other sites | 1.48 (1.3…1.68) | <0.001 |
| J00 | Acute nasopharyngitis [common cold] | 1.31 (1.25…1.37) | <0.001 |
| J01 | Acute sinusitis | 1.43 (1.34…1.52) | <0.001 |
| J06 | Acute upper respiratory infections of multiple and unspecified sites | 1.31 (1.24…1.38) | <0.001 |
| J11 | Influenza due to unidentified influenza virus | 1.38 (1.28…1.48) | <0.001 |
| J22 | Unspecified acute lower respiratory infection | 0.76 (0.64…0.9) | 0.002 |
| J30 | Vasomotor and allergic rhinitis | 1.45 (1.38…1.54) | <0.001 |
| J31 | Chronic rhinitis, nasopharyngitis and pharyngitis | 1.41 (1.32…1.5) | <0.001 |
| J32 | Chronic sinusitis | 1.48 (1.36…1.62) | <0.001 |
| J33 | Nasal polyp | 1.33 (1.2…1.47) | <0.001 |
| J34 | Other and unspecified disorders of nose and nasal sinuses | 1.55 (1.45…1.66) | <0.001 |
| J35 | Chronic diseases of tonsils and adenoids | 1.39 (1.3…1.49) | <0.001 |
| J37 | Chronic laryngitis and laryngotracheitis | 1.38 (1.26…1.51) | <0.001 |
| J38 | Diseases of vocal cords and larynx, not elsewhere classified | 1.53 (1.37…1.72) | <0.001 |
| J39 | Other diseases of upper respiratory tract | 1.45 (1.3…1.61) | <0.001 |
| J47 | Bronchiectasis | 1.46 (1.3…1.65) | <0.001 |
| J81 | Pulmonary oedema | 0.66 (0.52…0.83) | <0.001 |
| J91 | Pleural effusion, not elsewhere classified | 0.66 (0.61…0.7) | <0.001 |
| J96 | Respiratory failure, not elsewhere classified | 0.72 (0.65…0.79) | <0.001 |
| K00 | Disorders of tooth development and eruption | 1.5 (1.22…1.84) | <0.001 |
| K01 | Embedded and impacted teeth | 1.79 (1.65…1.95) | <0.001 |
| K02 | Dental caries | 1.31 (1.18…1.45) | <0.001 |
| K05 | Gingivitis and periodontal diseases | 1.35 (1.27…1.44) | <0.001 |
| K06 | Other disorders of gingiva and edentulous alveolar ridge | 1.44 (1.27…1.63) | <0.001 |
| K07 | Dentofacial anomalies [including malocclusion] | 1.65 (1.44…1.88) | <0.001 |
| K08 | Other disorders of teeth and supporting structures | 1.54 (1.42…1.67) | <0.001 |
| K09 | Other disorders of jaw | 1.34 (1.08…1.67) | 0.007 |
| K11 | Diseases of salivary glands | 1.39 (1.28…1.5) | <0.001 |
| K13 | Other diseases of lip and oral mucosa | 1.54 (1.43…1.67) | <0.001 |
| K14 | Diseases of tongue | 1.5 (1.34…1.67) | <0.001 |
| K21 | Gastro-esophageal reflux disease | 1.49 (1.39…1.59) | <0.001 |
| K30 | Functional dyspepsia | 1.47 (1.4…1.54) | <0.001 |
| K31 | Other diseases of stomach and duodenum | 1.54 (1.41…1.68) | <0.001 |
| K50 | Crohn disease [regional enteritis] | 1.53 (1.32…1.78) | <0.001 |
| K51 | Ulcerative colitis | 1.42 (1.3…1.56) | <0.001 |
| K57 | Diverticular disease of intestine | 1.36 (1.25…1.48) | <0.001 |
| K58 | Irritable bowel syndrome | 1.52 (1.44…1.61) | <0.001 |
| K60 | Fissure and fistula of anal and rectal regions | 1.42 (1.31…1.54) | <0.001 |
| K62 | Other diseases of anus and rectum | 1.44 (1.36…1.51) | <0.001 |
| K63 | Other diseases of intestine | 1.33 (1.22…1.45) | <0.001 |
| K70 | Alcoholic liver disease | 0.56 (0.49…0.65) | <0.001 |
| K86 | Other diseases of pancreas | 0.74 (0.66…0.82) | <0.001 |
| K90 | Intestinal malabsorption | 1.64 (1.52…1.78) | <0.001 |
| L00 | Staphylococcal scalded skin syndrome | 1.86 (1.52…2.28) | <0.001 |
| L01 | Impetigo | 1.3 (1.23…1.37) | <0.001 |
| L05 | Pilonidal cyst and sinus | 1.3 (1.11…1.53) | 0.001 |
| L13 | Other bullous disorders | 1.37 (1.07…1.75) | 0.01 |
| L21 | Seborrheic dermatitis | 1.47 (1.39…1.55) | <0.001 |
| L24 | Irritant contact dermatitis | 1.48 (1.41…1.56) | <0.001 |
| L25 | Unspecified contact dermatitis | 1.36 (1.3…1.42) | <0.001 |
| L28 | Lichen simplex chronicus and prurigo | 1.39 (1.31…1.47) | <0.001 |
| L29 | Pruritus | 1.5 (1.38…1.62) | <0.001 |
| L41 | Parapsoriasis | 1.74 (1.37…2.2) | <0.001 |
| L42 | Pityriasis rosea | 1.31 (1.23…1.39) | <0.001 |
| L43 | Lichen planus | 1.58 (1.47…1.69) | <0.001 |
| L50 | Urticaria | 1.39 (1.33…1.45) | <0.001 |
| L53 | Other erythematous conditions | 1.33 (1.19…1.49) | <0.001 |
| L56 | Other acute skin changes due to ultraviolet radiation | 1.49 (1.31…1.7) | <0.001 |
| L57 | Skin changes due to chronic exposure to ultraviolet radiation | 1.83 (1.62…2.08) | <0.001 |
| L60 | Nail disorders | 1.44 (1.39…1.49) | <0.001 |
| L62 | Disorders of hair in diseases classified elsewhere | 0.47 (0.37…0.61) | <0.001 |
| L64 | Androgenic alopecia | 1.63 (1.29…2.05) | <0.001 |
| L65 | Other nonscarring hair loss | 1.42 (1.23…1.65) | <0.001 |
| L68 | Hypertrichosis | 1.37 (1.11…1.68) | 0.003 |
| L70 | Acne | 1.58 (1.49…1.67) | <0.001 |
| L71 | Rosacea | 1.6 (1.51…1.7) | <0.001 |
| L72 | Follicular cysts of skin and subcutaneous tissue | 1.39 (1.32…1.47) | <0.001 |
| L73 | Other follicular disorders | 1.58 (1.45…1.73) | <0.001 |
| L80 | Vitiligo | 1.64 (1.39…1.94) | <0.001 |
| L81 | Other disorders of pigmentation | 1.9 (1.73…2.08) | <0.001 |
| L82 | Seborrheic keratosis | 1.97 (1.8…2.16) | <0.001 |
| L84 | Corns and callosities | 1.44 (1.36…1.52) | <0.001 |
| L85 | Other epidermal thickening | 1.61 (1.52…1.72) | <0.001 |
| L89 | Pressure ulcer | 0.64 (0.5…0.81) | <0.001 |
| L90 | Atrophic disorders of skin | 1.6 (1.5…1.72) | <0.001 |
| L91 | Hypertrophic disorders of skin | 1.6 (1.44…1.78) | <0.001 |
| L92 | Granulomatous disorders of skin and subcutaneous tissue | 1.43 (1.34…1.53) | <0.001 |
| L93 | Lupus erythematosus | 1.36 (1.15…1.61) | <0.001 |
| L94 | Other localized connective tissue disorders | 1.49 (1.34…1.67) | <0.001 |
| L98 | Other disorders of skin and subcutaneous tissue, not elsewhere classified | 1.31 (1.19…1.43) | <0.001 |
| M02 | Postinfective and reactive arthropathies | 1.52 (1.41…1.63) | <0.001 |
| M03 | Postinfective and reactive arthropathies in diseases classified elsewhere | 1.45 (1.02…2.06) | 0.04 |
| M05 | Rheumatoid arthritis with rheumatoid factor | 1.32 (1.24…1.4) | <0.001 |
| M06 | Other rheumatoid arthritis | 1.62 (1.5…1.74) | <0.001 |
| M07 | Enteropathic arthropathies | 1.55 (1.44…1.67) | <0.001 |
| M08 | Juvenile arthritis | 1.76 (1.46…2.11) | <0.001 |
| M11 | Other crystal arthropathies | 1.46 (1.25…1.71) | <0.001 |
| M16 | Osteoarthritis of hip | 1.34 (1.27…1.41) | <0.001 |
| M18 | Osteoarthritis of first carpometacarpal joint | 1.74 (1.46…2.07) | <0.001 |
| M20 | Acquired deformities of fingers and toes | 1.36 (1.27…1.46) | <0.001 |
| M21 | Other acquired deformities of limbs | 1.42 (1.3…1.54) | <0.001 |
| M22 | Disorder of patella | 1.56 (1.4…1.75) | <0.001 |
| M23 | Internal derangement of knee | 1.6 (1.47…1.75) | <0.001 |
| M24 | Other specific joint derangements | 1.34 (1.22…1.47) | <0.001 |
| M25 | Other joint disorder, not elsewhere classified | 1.37 (1.31…1.43) | <0.001 |
| M32 | Systemic lupus erythematosus | 1.34 (1.24…1.45) | <0.001 |
| M34 | Systemic sclerosis | 1.33 (1.08…1.62) | 0.006 |
| M35 | Other systemic involvement of connective tissue | 1.68 (1.53…1.85) | <0.001 |
| M45 | Ankylosing spondylitis | 1.58 (1.37…1.81) | <0.001 |
| M46 | Other inflammatory spondylopathies | 1.6 (1.48…1.73) | <0.001 |
| M47 | Spondylosis | 1.35 (1.25…1.46) | <0.001 |
| M50 | Cervical disc disorders | 1.37 (1.28…1.48) | <0.001 |
| M51 | Thoracic, thoracolumbar, and lumbosacral intervertebral disc disorders | 1.37 (1.25…1.5) | <0.001 |
| M62 | Other disorders of muscle | 1.4 (1.3…1.51) | <0.001 |
| M65 | Synovitis and tenosynovitis | 1.54 (1.4…1.68) | <0.001 |
| M66 | Spontaneous rupture of synovium and tendon | 1.68 (1.24…2.26) | <0.001 |
| M67 | Other disorders of synovium and tendon | 1.57 (1.41…1.74) | <0.001 |
| M68 | Disorders of muscle, ligament and fascia in diseases classified elsewhere | 1.58 (1.25…2) | <0.001 |
| M70 | Soft tissue disorders related to use, overuse and pressure | 1.46 (1.36…1.57) | <0.001 |
| M71 | Other bursopathies | 1.43 (1.33…1.55) | <0.001 |
| M72 | Fibroblastic disorders | 1.55 (1.45…1.66) | <0.001 |
| M75 | Shoulder lesions | 1.55 (1.42…1.7) | <0.001 |
| M76 | Enthesopathies of lower limb, excluding foot | 1.76 (1.61…1.93) | <0.001 |
| M77 | Other enthesopathies | 1.43 (1.34…1.52) | <0.001 |
| M79 | Other and unspecified soft tissue disorders, not elsewhere classified | 1.41 (1.37…1.46) | <0.001 |
| M81 | Osteoporosis without current pathological fracture | 1.52 (1.34…1.72) | <0.001 |
| M85 | Other disorders of bone density and structure | 1.42 (1.28…1.57) | <0.001 |
| M89 | Other disorders of bone | 1.47 (1.13…1.9) | 0.004 |
| M91 | Juvenile osteochondrosis of hip and pelvis | 1.49 (1.03…2.18) | 0.04 |
| M93 | Other osteochondropathies | 1.91 (1.7…2.13) | <0.001 |
| M94 | Other disorders of cartilage | 1.77 (1.58…1.99) | <0.001 |
| N02 | Recurrent and persistent haematuria | 1.31 (1.05…1.63) | 0.02 |
| N03 | Chronic nephritic syndrome | 1.48 (1.27…1.73) | <0.001 |
| N17 | Acute kidney failure | 0.7 (0.64…0.76) | <0.001 |
| N26 | Unspecified contracted kidney | 1.47 (1.03…2.1) | 0.04 |
| N31 | Neuromuscular dysfunction of bladder, not elsewhere classified | 1.41 (1.31…1.52) | <0.001 |
| N34 | Urethritis and urethral syndrome | 1.37 (1.24…1.51) | <0.001 |
| N35 | Urethral stricture | 1.35 (1.2…1.52) | <0.001 |
| N36 | Other disorders of urethra | 1.34 (1.12…1.61) | 0.001 |
| N40 | Benign prostatic hyperplasia | 1.45 (1.34…1.57) | <0.001 |
| N41 | Inflammatory diseases of prostate | 1.79 (1.67…1.92) | <0.001 |
| N42 | Other and unspecified disorders of prostate | 1.71 (1.57…1.86) | <0.001 |
| N43 | Hydrocele and spermatocele | 1.44 (1.33…1.56) | <0.001 |
| N45 | Orchitis and epididymitis | 1.48 (1.33…1.65) | <0.001 |
| N46 | Male infertility | 1.78 (1.72…1.84) | <0.001 |
| N47 | Disorders of prepuce | 1.46 (1.26…1.7) | <0.001 |
| N48 | Other disorders of penis | 1.88 (1.72…2.05) | <0.001 |
| N49 | Inflammatory disorders of male genital organs, not elsewhere classified | 1.57 (1.41…1.75) | <0.001 |
| N50 | Other and unspecified disorders of male genital organs | 2.18 (1.97…2.42) | <0.001 |
| N51 | Disorders of male genital organs in diseases classified elsewhere | 1.62 (1.48…1.76) | <0.001 |
| N62 | Hypertrophy of breast | 1.52 (1.38…1.67) | <0.001 |
| N63 | Unspecified lump in breast | 1.36 (1.27…1.45) | <0.001 |
| N64 | Other disorders of breast | 1.48 (1.28…1.72) | <0.001 |
| N74 | Female pelvic inflammatory disorders in diseases classified elsewhere | 1.67 (1.31…2.13) | <0.001 |
| N83 | Noninflammatory disorders of ovary, fallopian tube and broad ligament | 1.3 (1.15…1.46) | <0.001 |
| N89 | Other noninflammatory disorders of vagina | 1.37 (1.17…1.61) | <0.001 |
| N90 | Other noninflammatory disorders of vulva and perineum | 1.3 (1.19…1.42) | <0.001 |
| N91 | Absent, scanty and rare menstruation | 1.35 (1.14…1.61) | <0.001 |
| N95 | Menopausal and other perimenopausal disorders | 1.32 (1.15…1.52) | <0.001 |
| N96 | Recurrent pregnancy loss | 1.33 (1.14…1.56) | <0.001 |
| N97 | Female infertility | 1.33 (1.3…1.37) | <0.001 |
| N98 | Complications associated with artificial fertilization | 1.35 (1.15…1.59) | <0.001 |
| O28 | Abnormal findings on antenatal screening of mother | 1.51 (1.33…1.7) | <0.001 |
| O43 | Placental disorders | 1.32 (1.08…1.61) | 0.007 |
| O71 | Other obstetric trauma | 1.4 (1.29…1.52) | <0.001 |
| O87 | Venous complications and hemorrhoids in the puerperium | 1.47 (1.37…1.59) | <0.001 |
| Q05 | Spina bifida | 1.52 (1.22…1.9) | <0.001 |
| Q23 | Congenital malformations of aortic and mitral valves | 1.42 (1.25…1.62) | <0.001 |
| Q25 | Congenital malformations of great arteries | 1.41 (1.02…1.94) | 0.04 |
| Q53 | Undescended testicle | 2.24 (1.79…2.81) | <0.001 |
| Q61 | Cystic kidney disease | 1.65 (1.33…2.05) | <0.001 |
| Q65 | Congenital deformities of hip | 1.44 (1.19…1.73) | <0.001 |
| Q67 | Congenital musculoskeletal deformities of head, face, vertebral column and chest | 1.42 (1.22…1.66) | <0.001 |
| Q76 | Congenital malformations of spine and bony thorax, not elsewhere classified | 2.17 (1.36…3.46) | 0.001 |
| Q82 | Other congenital malformations of skin | 1.52 (1.35…1.72) | <0.001 |
| Q85 | Phacomatoses, not elsewhere classified | 1.71 (1.19…2.44) | 0.003 |
| Q87 | Other specified congenital malformation syndromes affecting multiple systems | 2.1 (1.4…3.13) | <0.001 |
| R00 | Abnormalities of heart beat | 1.35 (1.27…1.43) | <0.001 |
| R04 | Hemorrhage from respiratory passages | 1.3 (1.26…1.34) | <0.001 |
| R05 | Cough | 1.4 (1.34…1.47) | <0.001 |
| R06 | Abnormalities of breathing | 1.49 (1.39…1.6) | <0.001 |
| R07 | Pain in throat and chest | 1.39 (1.34…1.45) | <0.001 |
| R09 | Other symptoms and signs involving the circulatory and respiratory systems | 1.46 (1.02…2.09) | 0.04 |
| R12 | Heartburn | 1.34 (1.26…1.42) | <0.001 |
| R14 | Flatulence and related conditions | 1.49 (1.36…1.62) | <0.001 |
| R15 | Fecal incontinence | 1.69 (1.41…2.02) | <0.001 |
| R16 | Hepatomegaly and splenomegaly, not elsewhere classified | 0.74 (0.59…0.93) | 0.01 |
| R18 | Ascites | 0.64 (0.54…0.76) | <0.001 |
| R19 | Other symptoms and signs involving the digestive system and abdomen | 1.45 (1.36…1.56) | <0.001 |
| R20 | Disturbances of skin sensation | 1.49 (1.38…1.61) | <0.001 |
| R21 | Rash and other nonspecific skin eruption | 1.42 (1.33…1.52) | <0.001 |
| R22 | Localized swelling mass and lump of skin and subcutaneous tissue | 1.38 (1.32…1.44) | <0.001 |
| R23 | Other skin changes | 1.65 (1.48…1.84) | <0.001 |
| R25 | Abnormal involuntary movements | 1.36 (1.2…1.53) | <0.001 |
| R29 | Other symptoms and signs involving the nervous and musculoskeletal systems | 1.35 (1.23…1.47) | <0.001 |
| R30 | Pain associated with micturition | 1.57 (1.38…1.78) | <0.001 |
| R39 | Other and unspecified symptoms and signs involving the genitourinary system | 1.6 (1.44…1.77) | <0.001 |
| R42 | Dizziness and giddiness | 1.37 (1.31…1.44) | <0.001 |
| R43 | Disturbances of smell and taste | 1.38 (1.23…1.55) | <0.001 |
| R46 | Symptoms and signs involving appearance and behaviour | 1.4 (1.15…1.7) | <0.001 |
| R47 | Speech disturbances, not elsewhere classified | 0.76 (0.62…0.92) | 0.006 |
| R49 | Voice and resonance disorders | 1.76 (1.62…1.91) | <0.001 |
| R53 | Malaise and fatigue | 1.47 (1.38…1.58) | <0.001 |
| R54 | Senility | 0.62 (0.53…0.72) | <0.001 |
| R59 | Enlarged lymph nodes | 1.47 (1.34…1.61) | <0.001 |
| R61 | Generalized hyperhidrosis | 1.55 (1.44…1.66) | <0.001 |
| R64 | Cachexia | 0.73 (0.64…0.84) | <0.001 |
| R72 | Abnormality of white blood cells, not elsewhere classified | 1.72 (1.52…1.94) | <0.001 |
| R74 | Abnormal serum enzyme levels | 1.31 (1.21…1.42) | <0.001 |
| R76 | Abnormal immunological findings in serum | 1.34 (1.18…1.51) | <0.001 |
| R79 | Other abnormal findings of blood chemistry | 1.3 (1.06…1.59) | 0.01 |
| R82 | Other abnormal findings in urine | 1.35 (1.11…1.64) | 0.003 |
| R91 | Abnormal findings on diagnostic imaging of lung | 1.61 (1.39…1.87) | <0.001 |
| R93 | Abnormal findings on diagnostic imaging of other body structures | 2.02 (1.77…2.31) | <0.001 |
| R94 | Abnormal results of function studies | 1.58 (1.36…1.84) | <0.001 |
| S05 | Injury of eye and orbit | 1.32 (1.21…1.44) | <0.001 |
| S09 | Other and unspecified injuries of head | 1.3 (1.14…1.48) | <0.001 |
| S12 | Fracture of neck | 1.49 (1.15…1.93) | 0.002 |
| S23 | Dislocation, sprain and strain of joints and ligaments of thorax | 1.58 (1.18…2.11) | 0.002 |
| S33 | Dislocation and sprain of joints and ligaments of lumbar spine and pelvis | 1.32 (1.13…1.53) | <0.001 |
| S46 | Injury of muscle fascia and tendon at shoulder and upper arm level | 1.42 (1.31…1.53) | <0.001 |
| S56 | Injury of muscle and tendon at forearm level | 1.61 (1.06…2.45) | 0.03 |
| S69 | Other and unspecified injuries of wrist and hand | 1.36 (1.12…1.65) | 0.002 |
| S72 | Fracture of femur | 0.72 (0.61…0.85) | <0.001 |
| S76 | Injury of muscle fascia and tendon at hip and thigh level | 1.53 (1.39…1.68) | <0.001 |
| S83 | Dislocation and sprain of joints and ligaments of knee | 1.37 (1.29…1.44) | <0.001 |
| S86 | Injury of muscle fascia and tendon at lower leg level | 1.54 (1.41…1.68) | <0.001 |
| S96 | Injury of muscle and tendon at ankle and foot level | 1.4 (1.16…1.68) | <0.001 |
| S99 | Other and unspecified injuries of ankle and foot | 1.38 (1.06…1.82) | 0.02 |
| T02 | Fractures involving multiple body regions | 1.43 (1…2.05) | 0.05 |
| T17 | Foreign body in respiratory tract | 1.34 (1.17…1.54) | <0.001 |
| T20 | Burn and corrosion of head and neck | 1.76 (1.13…2.75) | 0.01 |
| T21 | Burn and corrosion of trunk | 1.34 (1.08…1.67) | 0.008 |
| T40 | Poisoning by, adverse effect of and underdosing of narcotics and psychodysleptics [hallucinogens] | 0.74 (0.55…0.99) | 0.04 |
| T51 | Toxic effect of alcohol | 0.59 (0.46…0.77) | <0.001 |
| T63 | Toxic effect of contact with venomous animals | 1.34 (1.24…1.44) | <0.001 |
| T75 | Effects of other external causes | 1.97 (1.51…2.57) | <0.001 |
| T78 | Adverse effects, not elsewhere classified | 1.43 (1.36…1.51) | <0.001 |
| T82 | Complications of cardiac and vascular prosthetic devices, implants and grafts | 1.36 (1.09…1.69) | 0.006 |
| T85 | Complications of internal prosthetic devices, implants and grafts, not elsewhere classified | 1.49 (1.2…1.85) | <0.001 |
| T86 | Complications of transplanted organs and tissue | 1.45 (1.16…1.82) | 0.001 |
| U11 | COVID-19 in screening | 1.56 (1.46…1.67) | <0.001 |
| Z01 | Encounter for other special examination without complaint suspected or reported diagnosis | 1.3 (1.24…1.36) | <0.001 |
| Z02 | Encounter for administrative examination | 1.3 (1.22…1.38) | <0.001 |
| Z03 | Encounter for medical observation for suspected diseases and conditions ruled out | 1.5 (1.44…1.56) | <0.001 |
| Z04 | Encounter for examination and observation for other reasons | 1.37 (1.29…1.45) | <0.001 |
| Z09 | Encounter for follow-up examination after completed treatment for conditions other than malignant... | 1.45 (1.38…1.53) | <0.001 |
| Z11 | Encounter for screening for infectious and parasitic diseases | 1.69 (1.44…2) | <0.001 |
| Z12 | Encounter for screening for malignant neoplasms | 1.36 (1.25…1.48) | <0.001 |
| Z13 | Encounter for screening for other diseases and disorders | 1.4 (1.29…1.52) | <0.001 |
| Z21 | Asymptomatic human immunodeficiency virus [HIV] infection status | 0.41 (0.28…0.59) | <0.001 |
| Z22 | Carrier of infectious disease | 1.36 (1.26…1.47) | <0.001 |
| Z23 | Encounter for immunization | 1.69 (1.62…1.77) | <0.001 |
| Z24 | Need for immunization against plague | 1.91 (1.77…2.05) | <0.001 |
| Z25 | Need for immunization against plague | 1.83 (1.74…1.92) | <0.001 |
| Z27 | Need for immunization against cholera | 1.8 (1.49…2.16) | <0.001 |
| Z28 | Immunization not carried out | 0.75 (0.65…0.86) | <0.001 |
| Z29 | Encounter for other prophylactic measures | 1.46 (1.34…1.59) | <0.001 |
| Z30 | Encounter for contraceptive management | 1.77 (1.45…2.15) | <0.001 |
| Z31 | Encounter for procreative management | 1.66 (1.53…1.8) | <0.001 |
| Z42 | Encounter for plastic and reconstructive surgery following medical procedure or healed injury | 1.53 (1.35…1.73) | <0.001 |
| Z46 | Encounter for fitting and adjustment of other prosthetic devices | 1.59 (1.45…1.75) | <0.001 |
| Z48 | Encounter for other postprocedural aftercare | 1.3 (1.25…1.36) | <0.001 |
| Z50 | Care involving use of rehabilitation procedures | 1.34 (1.08…1.67) | 0.009 |
| Z52 | Donors of organs and tissues | 1.83 (1.55…2.16) | <0.001 |
| Z56 | Problems related to employment and unemployment | 1.51 (1.35…1.69) | <0.001 |
| Z63 | Other problems related to primary support group, including family circumstances | 1.46 (1.31…1.64) | <0.001 |
| Z65 | Problems related to other psychosocial circumstances | 1.42 (1.01…2) | 0.04 |
| Z70 | Counseling related to sexual attitude behavior and orientation | 1.68 (1.54…1.83) | <0.001 |
| Z73 | Problems related to life management difficulty | 1.43 (1.28…1.6) | <0.001 |
| Z75 | Problems related to medical facilities and other health care | 1.46 (1.24…1.7) | <0.001 |
| Z80 | Family history of primary malignant neoplasm | 1.69 (1.52…1.89) | <0.001 |
| Z82 | Family history of certain disabilities and chronic diseases leading to disablement | 1.42 (1.23…1.64) | <0.001 |
| Z83 | Family history of other specific disorders | 1.8 (1.47…2.21) | <0.001 |
| Z86 | Personal history of certain chronic diseases, not elsewhere classified | 1.33 (1.19…1.48) | <0.001 |
| Z87 | Personal history of other diseases and conditions | 1.31 (1.23…1.39) | <0.001 |
| Z88 | Family history of allergy to drugs, medicaments and biological substances | 1.58 (1.18…2.12) | 0.002 |
| Z89 | Acquired absence of limb | 0.64 (0.53…0.77) | <0.001 |
| Z94 | Transplanted organ and tissue status | 1.38 (1.15…1.65) | <0.001 |
| Z98 | Other postprocedural states | 1.53 (1.43…1.62) | <0.001 |
